# Neoantigenic properties of *TP53* variants modify cancer risk in individuals with Li-Fraumeni syndrome

**DOI:** 10.1101/2025.06.24.25328545

**Authors:** Emilie Montellier, Olivier Manches, Jonathan Gaucher, Sandrine Blanchet, David Hoyos, Murielle Verboom, Christina M. Dutzmann, Sophie Coutant, Jacqueline Bou, Bertrand Fin, Robert Olaso, Jean-François Deleuze, Thierry Frebourg, Benjamin D. Greenbaum, Arnold J. Levine, Christian P. Kratz, Gaëlle Bougeard, Pierre Hainaut

## Abstract

**Importance:** Li-Fraumeni Syndrome (LFS) is an heterogenous cancer predisposition caused by pathogenic *TP53* variants, characterized by a lifelong high risk of a broad spectrum of cancers. At least certain pathogenic *TP53* variants have been shown be immunogenic in a somatic context. Whether neoantigenicity contributes to the heterogeneity of LFS is unknown.

**Objective:** To analyze the correlations between predicted neoantigenic properties of pathogenic *TP53* missense variants and LFS patterns.

**Design:** Association study between predicted MHC-I presentation scores for *TP53* hotspot variants, LFS presentation and individual HLA-I genotyping in carriers of *TP53* germline pathogenic variants using data from mutation databases and clinical registries.

**Setting:** MHC-I presentation scores were generated for the set of nonameric neo-peptides surrounding each *TP53* missense variant against 145 different HLA-I using NetMHCpan 4.1 and the Allele Frequency Net Database. Genotype-phenotype data were leveraged from the public *TP53* database (germline dataset, n=3,446; https://tp53.isb-cgc.org/) and two independent LFS clinical registries (n=339). Individual correlations between HLA-I genotyping, *TP53* missense variants and phenotypes were investigated in a group of 173 subjects with LFS.

**Participants:** Individuals with LFS enrolled in LFS-registries in France and Germany were included.

**Main outcomes and measures:** A predicted neoantigenic score (PNS) was calculated for each variant. Correlations with median age at first cancer, and cancer type were analyzed.

**Results:** Among 709 carriers of frequent *TP53* pathogenic variants, PNS was strongly correlated with median age at first cancer (range 18-43 years, p=0.01319, R=0.69). Compared to carriers of low PNS (<1) variants, carriers of high PNS (>2) variants showed delayed median age at first diagnosis (34 [CI95%, 29-40] vs. 25 years [CI95%, 22-27]), fewer sarcomas (osteosarcoma [RR 0.29, p-adj = 0.02]; soft-tissue [RR 0.41, p-adj = 0.02]), and more cancer types typically not associated with LFS [RR 1.61, p-adj = 0.02].

**Conclusion and Relevance:** MHC-I neoantigenic properties of *TP53* variants modify cancer risk and spectrum in carriers of pathogenic *TP53* variants, suggesting that individual variant-specific immune response may contribute to the heterogenous presentation of LFS.

## INTRODUCTION

Li-Fraumeni syndrome (LFS; Mendelian Inheritance in Man [MIM] #151623) is an autosomal dominant cancer predisposition syndrome associated with a high lifetime risk of several cancers caused by pathogenic germline (or postzygotic somatic mosaic) *TP53* variants, 80% of which are missense ^1^. Typically, LFS occurs in three phases, (1) a childhood phase (0-17 years), characterized by a quartet of early life cancers (adrenocortical carcinoma, ACC; choroid plexus tumor, CPT; medulloblastoma, MB, and rhabdomyosarcoma, RMS, all of which occur mostly before age ten years), and a high risk of other soft tissue sarcoma (STS), osteosarcoma (OS), and central nervous system (CNS) gliomas in adolescence; (2) an early adult phase (18-45 years), characterized by premenopausal breast cancer (PBC), multiple STS and glioma; and (3) a late adult phase (over 45 years), characterized by lung adenocarcinoma (LUAD), STS (typically leiomyosarcoma), colorectal cancer and prostate cancer ^2^. Clinical definitions are traditionally based on so-called “classic” LFS criteria ^3^, whereas revised Chompret criteria ^1,4,5^ capture the broad spectrum of LFS-related cancers and are aimed at identifying patients for *TP53* mutation testing.

The p53 protein is a multi-functional transcription factor regulating a complex network of cellular and systemic anti-proliferative responses ^6–8^. Loss of these functions, often caused by inactivating missense variants in the *TP53* gene, impairs several coordinated mechanisms of growth suppression that normally operate to counteract carcinogenesis ^9^. Missense variants in the *TP53* gene can have two major consequences on p53 protein expression: first they generate structural change that disrupts the interactions of the p53 protein with p53 DNA response elements with loss of p53 transcriptional activity. Second, mutant p53 proteins may escape Mdm2-dependent degradation and accumulate as stable proteins with dominant-negative or pro-oncogenic, gain of function effects ^10^.

In a recent systematic study, Hoyos et al. ^11^ have shown that the neoepitopes formed by different frequent somatic *TP53* variants could induce CD8+ T cells reactivity in peripheral blood mononuclear cells (PBMCs) from cancer patients as well as healthy donors. Furthermore, it was shown that predicted mutant p53 fitness was characterized by a trade-off between oncogenicity and immunogenicity with a selection of variants predicted to be poorly immunogenic. This correlation strongly supports that p53-driven immunoediting contributes to the selective pressures that shape cancer development and progression.

In this study we have investigated whether the predicted neoantigenic properties of *TP53* variants may contribute the heterogeneity of cancer phenotypes in carriers of germline *TP53* variants. First, we have used the prediction models of NetMHCpan 4.1 and the Allele Frequency Net Database ^12,13^ to extract immune recognition scores towards a panel of 145 HLA-class I antigens for a large dataset of *TP53* missense variants (n=2,314), including all missense variants reported to date in subjects and families matching LFS criteria. Second, we have analyzed the correlations between these predicted neoantigenic properties and cancer phenotypes (age at diagnosis, cancer types) in a large public dataset of germline *TP53* variant carriers ^14^ and in a validation cohort based on two clinical LFS registries (n=339) ^1,15^. Third, using genomics data from a subset of the patients of the validation cohort (n=173), we have examined the relationships between HLA-I-based variant neoantigenic prediction and tumor patterns at the individual level.

Our results show that variants with predicted strong neoantigenic properties were preferentially associated with attenuated presentations of LFS, suggesting that immune recognition of specific *TP53* variants may contribute to the individual variability of the syndrome.

## METHODS

### Predictions of TP53 variant neoantigenicity

Figure S1 shows a flowchart of the study. First, we generated a metrics of predicted neoantigenicity for a large dataset of 2,314 missense *TP53* variants for which yeast-assay based transcriptional activation scores were available ^16^. These scores were recently used to develop an integrative classification of missense variants in 4 classes according to a gradient of predicted severity (from class A, most severe, to class D, benign or likely benign) ^17^. Nine different, tiled 9-mer peptide sequences were generated *in silico* across each *TP53* missense variant and were analyzed using the NetMHCpan 4.1 ^12^ to extract their predicted affinity toward a large set of common HLA-I alleles (n = 145). For each missense variant, we produced 4 different metrics: (1) Minimal Affinity Score (MAS), corresponding to the lowest 9-mer peptide/HLA-I predicted affinity (in nanomol, nM), thus scoring the strongest predicted interaction for a given variant with any HLA-I. We considered affinity equal to or greater than 500 nM as non-relevant ^12^ and all variants with MAS above this threshold were assigned a value of 500 nM; (2): HLA Count Score (HCS) corresponding to the number of different HLA-I alleles with MAS <500 nM for each; (3) World Coverage Score (WCS), providing an estimate of the proportion of the world population with at least one HLA-I capable of interacting at MAS < 500 nM with the specified variant (based on population data from the Allele Frequency Net Database ^13^; (4) Amplitude Score (AMS), consisting in the ratio between MAS for the mutant peptide and the corresponding wild-type peptide. Three of these metrics (MAS, HCS, WCS) showed strong pairwise correlation (R>0.70, p-value <2.2e-16) and were interpolated to compute an integrated metric, the Predicted Neoantigenic Score (PNS). Each metric was normalized to a [0–1] scale. The affinity metric, for which lower values indicate stronger binding, was inverted prior to normalization. The PNS value was calculated as the sum of the three normalized metrics. This additive approach allowed each metric to contribute equally to the PNS. To analyze the correlations between PNS and cancer types in germline variant carriers, variants were aggregated in 3 PNS categories (low, PNS < 1; intermediate, PNS 1-2; high, PNS > 2).

### Correlations between TP53 genotypes, Neoantigenic Prediction Scores and LFS phenotypes

To examine whether PNS correlated with the severity of LFS phenotypes, we leveraged the NCI/IARC *TP53* germline database ^14^ (https://tp53.isb-cgc.org). Variants were classified in different pathogenicity classes based on structure-function analysis ^17^. We analyzed 144 class A variants in 1,426 carriers. A validation cohort was assembled, consisting of 339 carriers of class A variants from the French and German LFS registries ^1,15^. This validation cohort is maintained and annotated mainly independently of the NCI/IARC germline dataset.

### Individual correlations between TP53 genotype, HLA-I genotype, PNS and LFS phenotype

HLA-I genotypes were inferred from Whole Exome Sequencing data in a subgroup 173 patients from the validation cohort (**Table S2**). Patients were grouped in 4 categories according to the lowest MAS of their variant *TP53*-HLA-I pair: (1) ≤10 nM, considered as high affinity HLA-I binding, (2) >10 nM and <250 nM (intermediate), (3) over >250 nM (low), and (4) no relevant HLA-I binding.

### Graphical representation and statistical analysis

All graphics and statistical analyses were conducted using Rstudio. Spearman or Pearson correlation was performed to study relationship between variables. Pairwise Log-rank tests with a Bonferroni correction were performed used to assess differences in trend of cancer cumulative occurrence. Khi2 tests with Benjamini-Hochberg correction for multiple comparisons were performed to assess differences in proportion of tumor topologies. Multiple pairwise Wilcoxon tests with a Benjamini-Hochberg correction were used to compared median age of cancer diagnosis.

### Ethical approval and informed consent

Comprehensive genetics and clinical data were available for *TP53* variant carriers either identified at (1) the Genetics Department of Rouen University Hospital, with approval from CCTIRS (Comité Consultatif sur le Traitement de l’Information en matière de Recherche dans le domaine de la Santé) (N°07.291) and CNIL (Commission Nationale de l’Informatique et des Libertés) (N°907262), according to the French regulations, or (2) enrolled in the institutional review board–approved German Cancer Predisposition Syndrome Registry (DRKS00017382). Informed consent was obtained from all patients, parents, or legal guardians.

## RESULTS

Of the 2,314 variants analyzed, 1,185 had at least one 9-mer peptide predicted to bind at least one HLA-I allele with affinity <500 nM. **Table S1** lists, for each variant, the Predicted Neoantigenic Score (PNS) and each of the metrics on which it is based (MAS, HCS, WCS; see methods) and interacting HLA-I alleles. Overall, 144/145 tested HLA-I alleles were predicted to interact with at least one variant-derived peptide with a MAS< 500 nM (**Table S1**). Figure 1A shows the correlations between each of the metrics included in the PNS. We separated variants into 3 groups: Low (L, PNS <1), intermediate (I, PNS 1-2) and high (H, PNS >2). The distribution of the PNS showed a biphasic profile that correlated with variant functional class (Figure 1B), from D (more benign variants, lower PNS) to A (more severe variants, higher PNS; Chi-test p<0.001), consistent with the notion that more severe variants (e.g class A and B) corresponded to more disruptive and neoantigenic substitutions. Variants with I or H PNS tended to cluster within the DNA-binding and oligomerization domains of the p53 protein (Figure 1C). Frequent LFS variants, however, were mostly in the L PNS group. Only 10 variants had PNS ≥2.5, including 8 considered as severe (class A or B based on functional annotations ^17^). These variants were present in only 9/3446 (0.26%) carriers of the NCI/IARC germline *TP53* database. In contrast, the frequency of the common “hotspot” variants tended to be inversely correlated with the PNS score (Figure 1D). These observations are consistent with negative selection of variants with high predicted neoantigenicity among individuals with the clinical phenotype of LFS.

**Figure 1:**
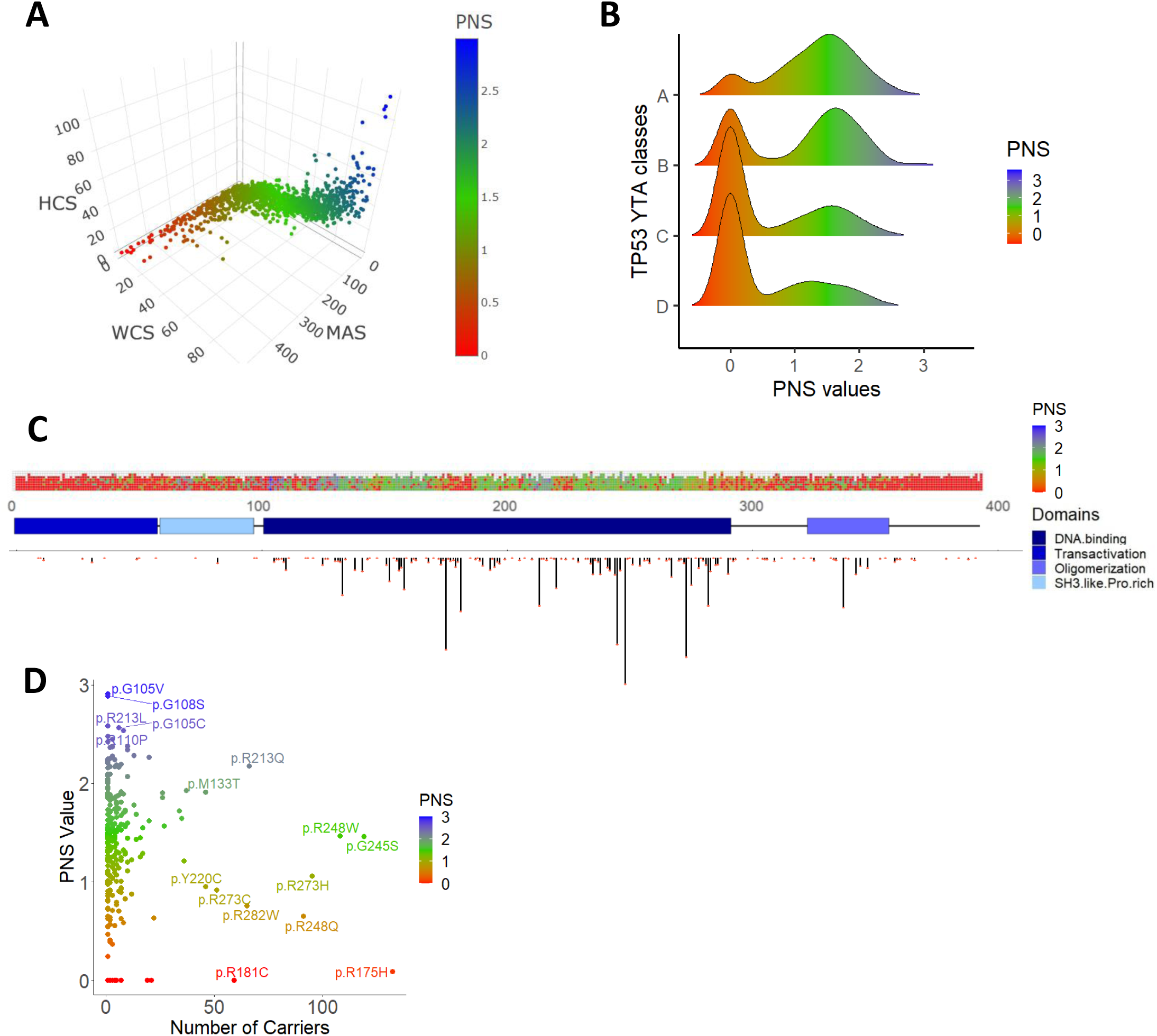
Mapping of the Predicted Neoantigenic Score (PNS) for *TP53*. **A.** Mapping of the Minimal Affinity Score (MAS), the World Coverage Score (WCS), and the HLA Count Score (HCS) for each of the 2,314 *TP53* misense variants. The 3 scores were normalized from 0 to 1 and an integrated score based on the sum of the 3 normalized scores was built (Predicted Neoantigenic Score, PNS, ranging from 0 to 3), and is represented with the color scale. **B.** Repartition of the PNS within the *TP53* variants classes (A, B, C and D). The density of PNS is displayed for each *TP53* variants class (YTA classes, from Montellier et al. 2024) **C.** Repartition of the PNS along p53 protein structure. Upper panel represents the PNS values at each p53 amino acid. The middle panel represents p53 secondary structure with codon numbers. The lower panel represents the count of germline *TP53* variant carriers at each amino acid (NCI/IARC germline database). **D.** Relationship between the number of germline *TP53* carriers and the PNS value for each *TP53* variant.

To evaluate the effect of predicted neoantigenicity on LFS presentation, we focused on class A variants, (n=402) which includes severely functionally impaired variants, consistent with Pathogenic (P) of Likely Pathogenic (LP) ClinVar annotations ^17^. Class A variants are present in 1426/3446 (41%) carriers of the NCI/IARC germline *TP53* database, including 865 carriers of 12 “hotspot” variants associated with severe LFS phenotypes, albeit with differences in cancer accrual over lifetime and in the median age at cancer diagnosis (from 19 years, (CI 95% [13-29]) for p.Y220C to 33 (CI 95% [22-42]) for p.C275Y, with p.R213Q as an outlier at median age 43 years (CI 95% [38-50])) (Figure 2**A,B,C**). Among these variants, 5 were scored as L PNS (p.R175H, p.R248Q, p.R282W, p.R273C, p.Y220C), 6 as I PNS (p.R273H, p.R337C, p.R248W, p.G245S, p.C275Y, p.R342P) and a single one 1 as H PNS (p.R213Q). Figure 2D shows a strong positive correlation between the median age at first cancer diagnosis and the PNS of these frequent *TP53* variants (R= 0.69, Pearson test p-value = 0.01319). Detailed analysis of the lifetime cancer accrual showed that, with increasing PNS, there was a progressive shift from early-life cancers (predominant in carriers of L PNS variants such as p.Y220C, p.R248Q, p.R282W, towards adult-life cancers (predominant in carriers of I/H PNS such as p.G245S, p.C275Y and p.R213Q carriers) (Figure 2C). Of note, each of the metrics integrated in the PNS (MAS, HCS, WCS) showed a significant correlation with age at cancer diagnosis (R>0.60, p<0.05). In contrast, AMS was not correlated with cancer latency (R=0.24, p=0.43) (**Figure S2**). Overall, these results support the notion that the predicted neoantigenic properties of *TP53* variants and their presentation by matched HLA-I are correlated with cancer latency and attenuation of LFS.

**Figure 2:**
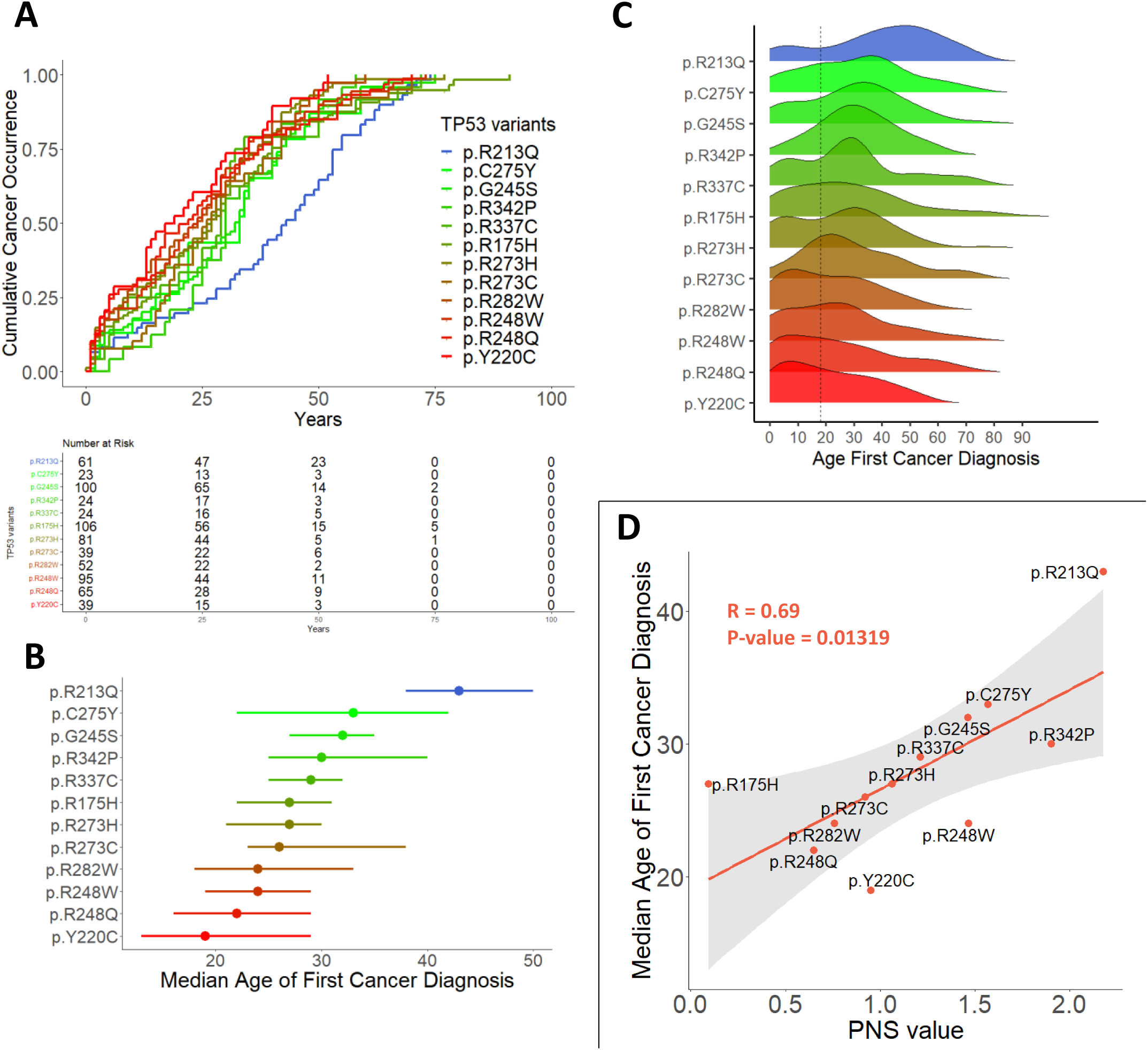
T*P*53 germline variants display variable cumulative cancer occurence patterns, which are correlated with the PNS. **A.** Variable cumulative cancer occurence patterns of frequent *TP53* variants from class A. Variants with at least 20 carriers were considered to conduct this analysis. The inverted Kaplan-Meier curves represent the age of diagnosis of the first cancer (n=709 individuals with age information, top panel). A Log-rank test was performed to assess differences between the curves (p-value = 0.00021). Number at risk table is displayed below the graph. **B.** Median age of first cancer diagnosis (dots) along with confidence interval at 95% (bars asides the dots) for each *TP53* variant. **C.** Pattern of age at first cancer diagnosis along lifetime. Density plot representing the pattern for each *TP53* variant. A vertical dotted line indicated age 18. **D.** Positive correlation of median age of first cancer diagnoses and the PNS value. For each frequent *TP53* variant, the median age of first cancer was plotted against the PNS value of the variant. The linear regression line is displayed in orange, and a Pearson correlation test was performed to assess relationship between the 2 variables (p-value = 0.01319, R = 0.69).

To examine which tumors of the LFS spectrum were the most affected by this attenuation, we analyzed the phenotypes of all class A variant carriers (n=1,426), irrespective of the variant’s hotspot status. Figure 3A shows that PNS predicted significant differences in cancer latency, with a 9 years delay in the median age at cancer diagnosis between carriers of H PNS (34 years [CI95%, 29-40], vs L PNS variants (25 years [CI95%, 22-27]); pairwise Log-rank test p-adj = 0.00088) and an intermediate median age in carriers of I PNS variants (29 years [CI95%, 27-30]; pairwise Log-rank test p-adj = 0.00191). Attenuation was not uniformly observed across all cancers of the LFS spectrum. A significant decrease was observed in carriers of H PNS versus L PNS variants in the proportion of osteosarcoma [RR 0.29, p-adj = 0.02] and soft tissues sarcoma [RR 0.41, p-adj = 0.02], as well as tendency for decreased proportion of hematopoietic malignancies. In contrast, cancers considered as not characteristic of the LFS spectrum were increased [1.61, p-adj = 0.02]. PNS did not appear to have a significant effect on the proportion of breast, brain cancers or adrenal cortical carcinomas, although a tendency for increased proportion was consistently observed for the latter cancer (Figure 3B). The age distribution of most cancers also varied according to PNS. Significant differences in the median age at diagnosis were observed for breast cancer (H PNS vs. L PNS, 39 vs. 31 years, p-adj = 0,00056; H PNS vs. I PNS, 39 vs. 32 years, p-adj = 0,00089) and non-LFS-spectrum cancers (H PNS vs. L PNS, 49.5 vs. 36 years, p-adj = 0,0000058; H PNS vs. I PNS, 49.5 vs. 36 years, p-adj = 0,0000058). Variants with higher PNS values were also associated a trend for increased latency of adrenal cortical carcinomas and of hematopoietic malignancies, whereas the age distribution of osteosarcoma, soft tissues sarcoma and brain tumors were similar across the 3 PNS groups (Figure 3C).

**Figure 3:**
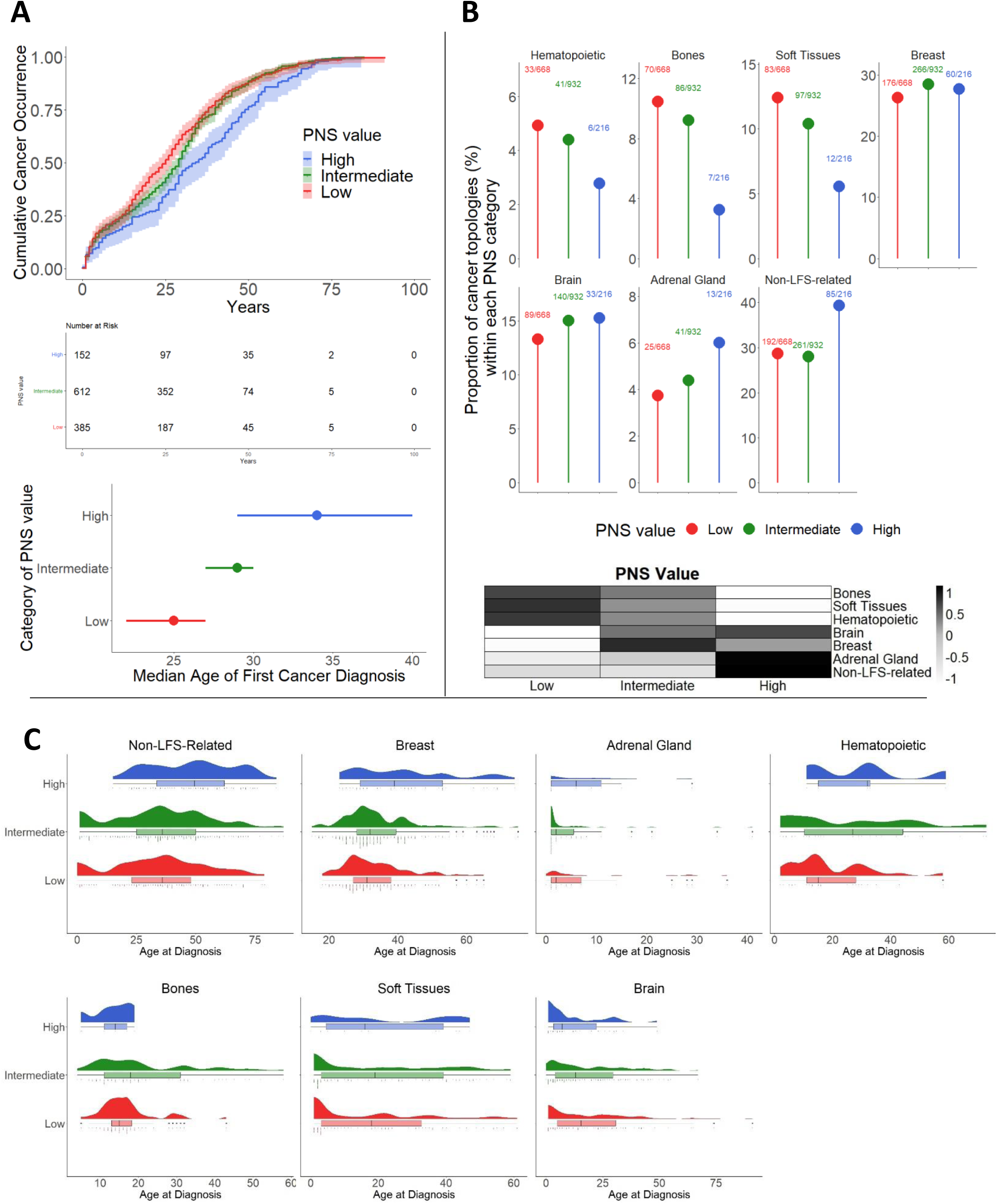
Strength of PNS value is associated with an attenuated LFS phenotype. **A.** Cumulative cancer occurence patterns vary according to the PNS category. Individuals from NCI/IARC germline *TP53* database carrying a class A variant (n=1,426) were separated into 3 groups based on the PNS value of the *TP53* variants, irrespective of the variant’s hotspot status. PNS categories: High for PNS >2, Intermediate for 1≤ PNS ≤2, and Low for PNS <1. The inverted Kaplan-Meier curves represent the age of first cancer diagnosis for each individual with age information (1,149 individuals, top panel). A Log-rank test was performed to assess differences between the curves (p-value = 0.00079). Pairwise Log-rank tests were performed to assess comparisons between PNS groups with a Bonferroni correction: low vs. high p-adj = 0.00088; high vs. intermediate p-adj = 0.00191. Number at risk table is displayed below the graph. The bottom panel represents the median age of first cancer diagnosis (dots) along with confidence interval at 95% (bars asides the dots). **B.** Distribution of cancers’ topology vary according to the PNS category. The typical LFS topologies are displayed (hematopoietic system, bones, soft tissues, breast, brain and adrenal gland) and non-LFS-spectrum topologies are grouped. For each cancer topology, the ratio of number of cases (n) over the total number of cancers analyzed within each PNS value group (N) is indicated on the top of the dot representing the percentage of cancer topology. Total number of cancer analyzed n=1,816. Multiple pairwise comparisons of proportion of topologies between the different PNS categories are performed using logistic model based on Khi2 statistic to extract risk ratio, confidence intervals at 95%, and an adjusted p-value (correction for multiple comparisons using Benjamini-Hochberg method). Significant risk ratio High vs. Low PNS: Bones [RR 0.29, p-adj = 0.02], Soft Tissues [RR 0.41, p-adj = 0.02], Non-LFS-spectrum [1.61, p-adj = 0.02]. Heatmap (bottom panel) synthetizes cancer topology distribution from top panel (normalization in row by cancer topologies, and color scale represents enrichment scores. **C.** Age distributions of cancers of different topologies vary according to the PNS category. The rain-cloud plots display (1) a density plot showing distribution of age of diagnosis of cancers, (2) a box plot showing median age of diagnosis as well as quartiles and outlier, and (3) a dot plot showing every cancer analyzed. Multiple pairwise comparison of median age of cancer is performed between PNS groups for each cancer topology by running Wilcoxon tests with a Benjamini-Hochberg correction. Significant differences has been determined for breast cancer (High vs. Low, age 39 vs. 31, p-adj = 0,00056; High vs. Intermediate, age 39 vs. 32, p-adj = 0,00089) and non-LFS-spectrum cancers (High vs. Low, age 49.5 vs. 36, p-adj = 0,0000058; High vs. Intermediate, age 49.5 vs. 36, p-adj = 0,0000058).

Overall, these results suggest that PNS correlated with selective attenuation of the LFS phenotype. Interestingly, this attenuation recapitulated the phenotypic characteristics of carriers of class B or C variants, who tended to present with less severe and more heterogeneous LFS phenotypes than class A variant carriers^17^. Indeed, cumulative cancer occurrence in carriers of H PNS class A variants was similar to the one observed in carriers of class B variants (**Figure S4A**). Likewise, the shifts in cancer patterns observed between carriers of L and H PNS class A variants resembled those previously reported between carriers of class A versus class B or C patients (**Figure S4B**). Limited numbers of carriers and heterogeneous phenotypes precluded the statistical analysis of the correlations between PNS and of cancer phenotypes in class B and C carriers.

These analyses were repeated in a validation cohort consisting of LFS patient carrying class A variant in the French and German clinical LFS registries (n=339) ^1,15^. This analysis confirmed the significant differences in lifetime cancer accrual between carriers of H versus L PNS class A variants (median age at first cancer diagnosis: 31 years [CI 95% 24-38] in H PNS vs. 25 years [CI95% 21-28] in I PNS and 23 years [CI95% 17-27] in L PNS variant carriers, respectively; Log-Rank test, p-value = 0.03) (**Figure S3A**). The validation cohort recapitulated the trends in tumor patterns observed in carriers of the NCI/IARC germline *TP53* database, although small numbers precluded statistical significance. H PNS variant carriers presented with a lower proportion of osteosarcomas (2.2% vs. 7.3% and 12.0%), soft tissue sarcomas (11.1% vs. 14.0% and 16.9%) and hematological malignancies (2.2% vs. 6.1% and 7.0%) compared L and I PNS carriers (**Figure S2B**). In contrast, they presented with a higher proportion of adrenal cortical carcinoma (15.6% vs. 8.5% and 6.3%) and cancers considered as not characteristic of the LFS spectrum (22.2 vs. 14.0% and 12.0%) (**Figure S2B**).

To address whether a protective effect of neoantigenic variant prediction could be observed at individual levels, we used HLA-I genotyping data from 173 patients of the validation cohort ^1,15^ (Figure 4). Patients were stratified in 4 groups according to the Minimal Affinity Score (MAS) of their most effective HLA-I/variant peptide pair (High, MAS ≤10 nM, Intermediate, MAS>10 and ≤250 nM, Low, MAS>250 and ≤500 nM, Negative, no identified HLA at MAS ≤500 nM). Only 4 (2%) were in the High group, whereas 40 (25%) and 17 (10%) were in the Intermediate and Low groups, respectively and 122 (65%) were in the Negative group. The cancer spectrum differed between the groups. The Negative and Low groups presented with the most severe phenotypes, recapitulating the broad LFS spectrum dominated by cancers of the adrenal glands, brain, bones, soft tissues, breast and hematopoietic system, or with non-LFS spectrum cancers. The Intermediate and, in particular, the High group presented with an attenuated LFS phenotype, with an increased proportion or carriers who are cancer-free or presenting cancers not typical of the LFS spectrum and a reduced proportion of cancers of the bones and soft tissues. In particular, patients of the high group developed only adrenal cortical carcinoma (1 case) or breast cancer (2 cases), whereas 1 was cancer-free. These results show that carriage of high-affinity HLA was infrequent in this cohort of patients selected for LFS phenotype. However, when present, intermediate or high affinity HLA were consistent with a tumor type-specific phenotypic attenuation which recapitulated the observations on carriers of the NCI/IARC germline *TP53* database and of the validation cohort.

**Figure 4:**
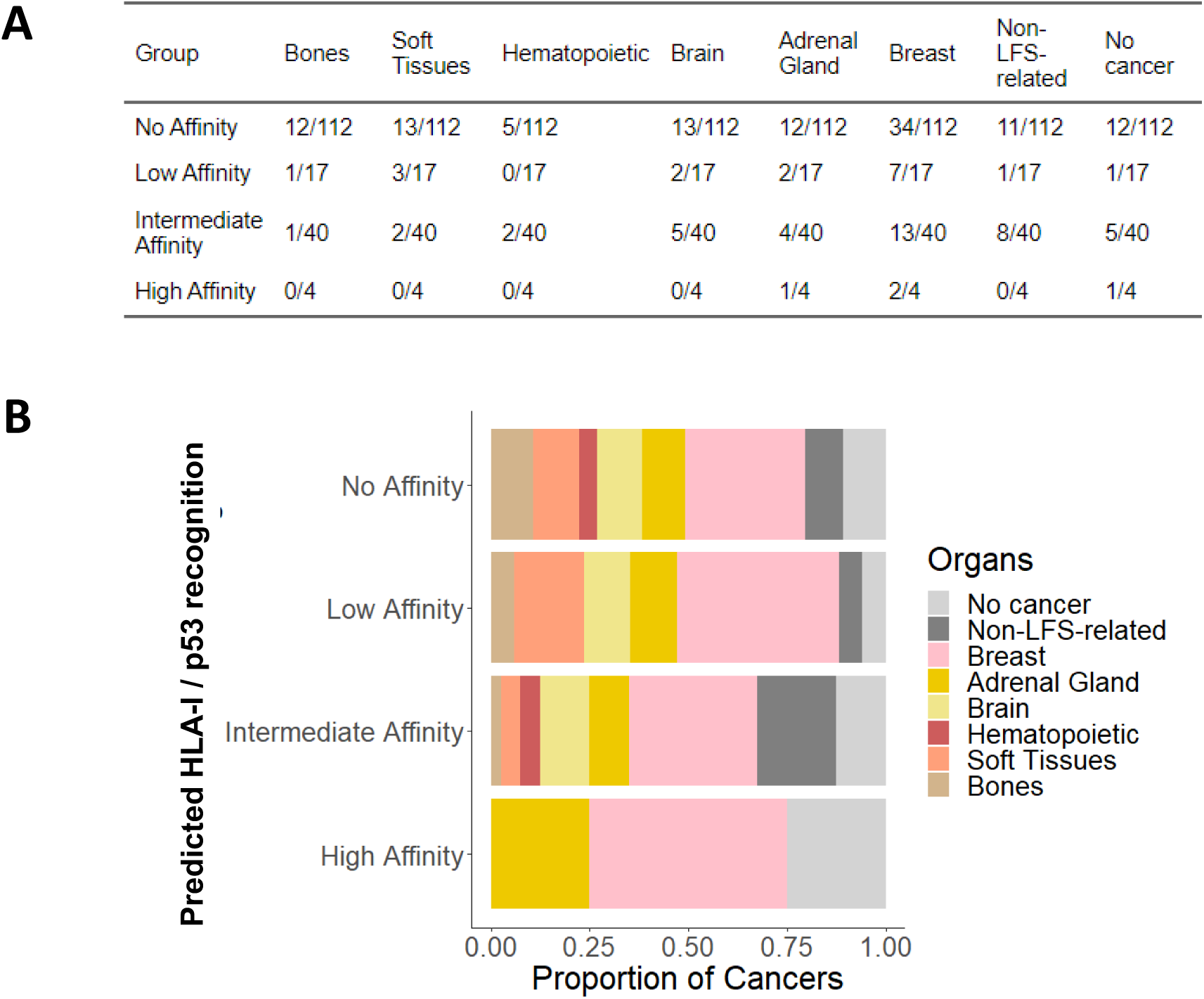
Strong predicted neoantigenic response determined by individual HLA-I genotyping of LFS patients correlates with attenuated LFS phenotype. **A.** Repartition of cancer topologies within each group of predicted neoantigenic response. The 6HLA-I alleles types combined with the *TP53* variant identity were used to give an individual score for each patient, corresponding to the strongest affinity of any of his 6 HLA-I for the *TP53* variant carried. LFS patients (n=173) were separated into 4 categories: high for affinity ≤ 10 nM, intermediate for 10 nM > affinity ≤ 250 nM, low for 250 nM > affinity < 500 nM, and no affinity when no HLA-I is predicted to interact with the variant. First cancers are analyzed (n=169). **B.** The proportions of cancer topologies vary among groups of LFS patients separated based on the predicted neoantigenic response.

## DISCUSSION

Our results show that the neoantigenic properties of p53 protein variants modify the cancer risk in individuals with LFS. The presented data suggest that pathogenic *TP53* germline variants can elicit a neoantigenic immune response that delays the occurrence and selectively attenuates cancers of the LFS spectrum. Thus, difference in neoantigenicity between variants and among individual patients may contribute to the broad clinical heterogeneity observed among individuals with LFS. Our results confirm and extend to germline variants the fitness selection model proposed by Hoyos et al. ^11^ for somatic *TP53* variants, suggesting that cancer-associated variants are selected to optimally solve an evolutionary trade-off between oncogenic potential and neoantigen immunogenicity. Whereas severe variants (class A) tend to have greater neoantigenic qualities than benign ones (class D), the most commonly represented “hotspots” LFS variants correspond to gaps in the HLA-I neoantigenic repertoire, enabling these variants to escape immune recognition and cause severe LFS phenotypes. Thus, the *TP53* variant spectrum observed in individuals with LFS reflects the combined effects of preferential mutagenesis, positive selection of severely impaired protein variants and negative selection of highly neoantigenic variants.

Whereas there is strong evidence that somatic *TP53* variants can elicit such a sustained and specific T-cell response, this has not yet been demonstrated in germline *TP53* mutation carriers. A key question is to determine how neoantigenic variants escape central or peripheral tolerance during development. Our results are consistent with the notion that, under normal circumstances, p53 protein levels, either wild-type or mutant, are tightly regulated and kept at very low levels during development and early life, only rising during stress or oncogenic transformation. Whereas LFS tumors often show stabilized and overexpressed mutant p53, whether this accumulation takes place in normal LFS cells and tissues is still a matter of debate ^18,19^. In a recent study of LFS patients-derived fibroblasts expressing p.R248Q, Agarwal et al. ^20^ have shown that this pathogenic mutant variant only accumulated after loss of the wild-type allele (LOH) in a sub-population of cells that showed delayed senescence, increased DNA damage and enhanced metabolic activity, conferring them a competitive advantage that drives their clonal expansion within the culture. Thus, LOH may constitute the critical event causing mutant p53 to become exposed and available for neoantigenic presentation and the variant may therefore be seen by the immune system as “somatic”. Consistent with this interpretation, we noted that, among the predictors of neoantigenicity generated by NetMHCpan 4.1, Amplitude, a metrics that evaluates the difference in affinity between a neoantigen and the corresponding wild-type peptide, was only poorly correlated with age at cancer onset, in contrast with other predictors (**Figure S2**). This indicates that the difference in HLA-I binding between wild-type and mutant peptides is not a major determinant of its neoantigenicity, suggesting that these peptides induce no or only incomplete central tolerance during development.

Our results show that immune attenuation does not equally affect all types of cancers within the LFS spectrum. Whereas high PNS variants appeared to be associated with delayed onset of most cancers, cancer risk was significantly reduced only for osteosarcoma, soft-tissue sarcoma. In contrast, the risk of adrenal cortical carcinoma and breast cancer appeared to remain relatively unaffected. These differences might be caused by several mechanisms, including immune privilege and restricted access to specific types of cancers, default of certain cancers in processing the relevant neoantigenic peptides, or the need for tissue-specific additional signals to trigger full T-cell activation. Mutant p53 also causes profound alterations of immune regulations with documented effects on macrophages, dendritic cells, natural killer cells (NK), T cells, and B cells ^21^. Such effects may enable certain cancers to escape immune surveillance despite the presence of a potent neoantigen. An alternative explanation is that neoantigenic attenuation is inversely proportional to the number of LFS cancer initiating events occurring in different tissues. Indeed, the phenotypic attenuation caused by variant neoantigenicity is remarkably similar to the one caused by variant functionality. In other words, tumor phenotypes in carriers of class A (most severe) variants with high or intermediate PNS are very similar to those of carriers of class B or C (less severe) variants with low PNS. A realistic hypothesis is that class B or C variants may support the emergence of less tissue-specific LFS cancer-initiating cells than class A variants. Thus, neoantigenic attenuation may similarly operate by controlling the pools of such initiating cells, causing a significant risk reduction in tissues where these pools are rate-limiting, but not in tissues in which a high number of initiating event enables the persistence of sufficient pools even in the face of immune attenuation. As a result, the cancers that appear the less sensitive to neoantigenic attenuation are precisely those that are the most frequent in children and adult LFS patients, namely, adrenal cortical and breast cancers, respectively.

These results have implications for predicting cancer risk and surveillance in germline *TP53* mutation carriers. As shown in our analysis of tumor patterns in carriers with known HLA-I alleles (Figure 4), the vast majority to patients with severe LFS phenotypes appear to lack high-affinity HLAs matched to their variants. HLA typing may however provide an explanatory variable in subjects who present attenuated, atypical or even no LFS phenotype despite carriage of a pathogenic/likely pathogenic variant. In this respect, neoantigenicity may contribute to inter-individual and intra-familial heterogeneity in LFS presentation. In the long term, and after due evaluation in a clinical context, HLA typing may thus contribute to determine improved and personalized risk assessment and surveillance strategies. On the other hand, it may also become possible to exploit variant neoantigenicity in individualized immuno preventive and immunotherapeutic approaches.

The limitations of this work are, first and foremost, its correlative nature. Whereas it reveals a strong correlation between LFS latency, risk and variant neoantigenicity, it does not demonstrate the existence of an effective cancer protective or suppressive anti-mutant p53 T cell response in germline *TP53* variant carriers. Further experimental and clinical studies are mandatory to evaluate the precise extend, dynamics and suppressive efficacy of such an immune response. Another limitation is that it is based on predicted interactions between HLA-I and every possible 9-mer peptide derived from missense *TP53* variants, without taking into account which peptides may actually be effectively cleaved and processed by the immunoproteasome and by cell-specific intermediate proteasomes. In our analysis, the usage of current standard peptide cleavage prediction models proved to restrict the set of peptides deemed as potential neoantigens and did not increase the accuracy of phenotypic correlations, suggesting that these models may not adequately capture the diversity of peptide proteolytic mechanisms that lead to the recognition of p53 protein variants.

## CONCLUSION

Predicted neoantigenic qualities of pathogenic *TP53* germline variants are correlated with variant frequency and cancer latency in germline variant carriers. This supports that variants can elicit a T-cell immune response contributing to the attenuation of the Li-Fraumeni Syndrome. Variant neoantigenic prediction scores may help in assessing cancer risk in carriers of pathogenic *TP53* variants with attenuated LFS phenotypes.

## Data Availability

All data produced in the present study are available upon reasonable request to the authors.

## SUPPLEMENTARY FIGURES LEGEND

**Figure S1**: Flowchart of the study: generation of the Predicted Neoantigenic Score (PNS) for *TP53* variants and subsequent genotype / phenotype correlations in germline *TP53* carriers.

**Figure S2**: Correlation of individual neoantigenic scores with median age of first cancer diagnosis for the 12 *TP53* class A hotspots (supplemental information related to Figure 2D). Correlation of median age of first cancer diagnoses and the 3 scores used to build PNS value (MAS, HCS and WCS), whereas no correlation exists with amplitude score (AMS). For each frequent *TP53* variant, the median age of first cancer was plotted against the individual neoantigenic scores. The linear regression line is displayed in orange, and a Spearman correlation test was performed to assess relationship between the variables (MAS: p=0.0276, R = -0.63; HCS, p=0.01683, R = 0.67; WCS p=0.03367, R = 0.61; AMS p=0,4349, R = 0.24).

**Figure S3**: Strenght of PNS value is associated with an attenuated LFS phenotype in a validation cohort of LFS patients. **A.** Cumulative cancer occurrence patterns vary according to the PNS category. Individuals from the validation cohort (LFS families), carrying a class A variant (n = 339 including no cancer individuals) were separated into 3 groups based on the PNS value of the *TP53* variants. PNS categories: High for PNS >2, Intermediate for 1≤ PNS ≤2, and Low for PNS <1. The inverted Kaplan-Meier curves represent the age of diagnosis of the first cancer for each individual (top panel). A Log-rank test was performed to assess differences between the curves (p = 0.03). Number at risk table is displayed below the graph. The bottom panel represents the median age of first cancer diagnosis (dots) along with confidence interval at 95% (bars asides the dots). **B.** Distribution of cancers’ topologies vary according to the PNS category. The typical LFS topologies are displayed (hematopoietic system, bones, soft tissues, breast, brain and adrenal gland) and non-LFS-spectrum topologies are grouped. First cancer(s) of patients are analyzed (n=351). For each cancer topology, the ratio of number of cases (n) over the total number of cancers analyzed within each PNS value group (N) is indicated on the top of the dot representing the percentage of cancer topology.

**Figure S4**: Class A LFS patients carrying a *TP53* variant with high PNS have shared traits with class B and C carriers (supplementary information related to Figure 3). **A.** Cumulative cancer occurrence of class B, C and D individuals compared with class A separated into high, intermediate and low PNS. Individuals from NCI/IARC germline *TP53* database carrying a single missense variant (n=2,129) were separated into class A, B, C and D. Class A were sub classified into 3 groups based on the PNS. PNS categories: High for PNS >2, Intermediate for 1-2, and Low for PNS <1. The inverted Kaplan-Meier curves represent the age of first cancer diagnosis for each individual with age information (1,665 individuals, top left panel). Number at risk table is displayed below the graph. A Log-rank test was performed to assess differences between the curves (p-value <0.0001). Pairwise Log-rank tests were performed to assess comparisons between classes and PNS groups with a Bonferroni correction (right panel). **B.** Distribution of cancers’ topology vary according to the classes and PNS categories. The typical LFS topologies are displayed (hematopoietic system, bones, soft tissues, breast, brain and adrenal gland) and non-LFS-spectrum topologies are grouped. Total number of cancer analyzed n=2,526. **C.** Age distributions of cancers of different topologies vary according to the classes and PNS categories. The rain-cloud plots display (1) a density plot showing distribution of age of diagnosis of cancers, (2) a box plot showing median age of diagnosis as well as quartiles and outlier, and (3) a dot plot showing every cancer analyzed.

**Table S1**: 2,314 *TP53* missense variants along with neoantigenic scores (related to Figure 1)

**Table S2**: List of the 145 HLA-I studied and the number of the predicted *TP53* variants recognized (related to Figure 1)

**Table S3**: Dataset of LFS patients with *TP53* variant identity, HLA-I typing, and clinical data (related to Figure 4)

## BIBLIOGRAPHY

1. Bougeard G, Renaux-Petel M, Flaman JM, et al. Revisiting Li-Fraumeni Syndrome From TP53 Mutation Carriers. J Clin Oncol. 2015;33(21):2345–2352. doi:10.1200/JCO.2014.59.5728

2. Amadou A, Achatz MIW, Hainaut P. Revisiting tumor patterns and penetrance in germline TP53 mutation carriers: temporal phases of Li-Fraumeni syndrome. Curr Opin Oncol. 2018;30(1):23–29. doi:10.1097/CCO.0000000000000423

3. Li FP, Fraumeni JF, Mulvihill JJ, et al. A cancer family syndrome in twenty-four kindreds. Cancer Res. 1988;48(18):5358–5362.

4. Frebourg T, Bajalica Lagercrantz S, Oliveira C, Magenheim R, Evans DG, European Reference Network GENTURIS. Guidelines for the Li-Fraumeni and heritable TP53-related cancer syndromes. Eur J Hum Genet. 2020;28(10):1379-1386. doi:10.1038/s41431-020-0638-4

5. Saucier E, Bougeard G, Gomez-Mascard A, et al. Li-Fraumeni-associated osteosarcomas: The French experience. Pediatr Blood Cancer. 2024;71(12):e31362. doi:10.1002/pbc.31362

6. Kastenhuber ER, Lowe SW. Putting p53 in Context. Cell. 2017;170(6):1062–1078. doi:10.1016/j.cell.2017.08.028

7. Kruiswijk F, Labuschagne CF, Vousden KH. p53 in survival, death and metabolic health: a lifeguard with a licence to kill. Nat Rev Mol Cell Biol. 2015;16(7):393–405. doi:10.1038/nrm4007

8. Levine AJ. p53: 800 million years of evolution and 40 years of discovery. Nat Rev Cancer. 2020;20(8):471–480. doi:10.1038/s41568-020-0262-1

9. Hainaut P, Pfeifer GP. Somatic TP53 Mutations in the Era of Genome Sequencing. Cold Spring Harb Perspect Med. 2016;6(11):a026179. doi:10.1101/cshperspect.a026179

10. Wang J, Liu W, Zhang L, Zhang J. Targeting mutant p53 stabilization for cancer therapy. Front Pharmacol. 2023;14:1215995. doi:10.3389/fphar.2023.1215995

11. Hoyos D, Zappasodi R, Schulze I, et al. Fundamental immune–oncogenicity trade-offs define driver mutation fitness. Nature. 2022;606(7912):172–179. doi:10.1038/s41586-022-04696-z

12. Reynisson B, Alvarez B, Paul S, Peters B, Nielsen M. NetMHCpan-4.1 and NetMHCIIpan-4.0: improved predictions of MHC antigen presentation by concurrent motif deconvolution and integration of MS MHC eluted ligand data. Nucleic Acids Res. 2020;48(W1):W449–W454. doi:10.1093/nar/gkaa379

13. Gonzalez-Galarza FF, McCabe A, Santos EJMD, et al. Allele frequency net database (AFND) 2020 update: gold-standard data classification, open access genotype data and new query tools. Nucleic Acids Res. 2020;48(D1):D783–D788. doi:10.1093/nar/gkz1029

14. de Andrade KC, Lee EE, Tookmanian EM, et al. The TP53 Database: transition from the International Agency for Research on Cancer to the US National Cancer Institute. Cell Death Differ. 2022;29(5):1071–1073. doi:10.1038/s41418-022-00976-3

15. Penkert J, Strüwe FJ, Dutzmann CM, et al. Genotype-phenotype associations within the Li-Fraumeni spectrum: a report from the German Registry. J Hematol Oncol. 2022;15(1):107. doi:10.1186/s13045-022-01332-1

16. Kato S, Han SY, Liu W, et al. Understanding the function-structure and function-mutation relationships of p53 tumor suppressor protein by high-resolution missense mutation analysis. Proc Natl Acad Sci U S A. 2003;100(14):8424–8429. doi:10.1073/pnas.1431692100

17. Montellier E, Lemonnier N, Penkert J, et al. Clustering of TP53 variants into functional classes correlates with cancer risk and identifies different phenotypes of Li-Fraumeni syndrome. iScience. 2024;27(12). doi:10.1016/j.isci.2024.111296

18. Kuba MG, Lester SC, Bowman T, et al. Histopathologic features of breast cancer in Li-Fraumeni syndrome. Mod Pathol. 2021;34(3):542–548. doi:10.1038/s41379-020-0610-4

19. MacGeoch C, Turner G, Bobrow LG, Barnes DM, Bishop DT, Spurr NK. Heterogeneity in Li-Fraumeni families: p53 mutation analysis and immunohistochemical staining. J Med Genet. 1995;32(3):186-190. doi:10.1136/jmg.32.3.186

20. Agarwal H, Tal P, Goldfinger N, et al. Mutant p53 reactivation restricts the protumorigenic consequences of wild type p53 loss of heterozygosity in Li-Fraumeni syndrome patient-derived fibroblasts. Cell Death Differ. 2024;31(7):855–867. doi:10.1038/s41418-024-01307-4

21. Levine AJ. P53 and The Immune Response: 40 Years of Exploration—A Plan for the Future. Int J Mol Sci. 2020;21(2):541. doi:10.3390/ijms21020541

