## Supplementary Figures for "Neoantigenic properties of *TP53* variants modify cancer risk in individuals with Li-Fraumeni syndrome"

(1) Univ. Grenoble Alpes, Inserm 1209, CNRS 5309, Institute for Advanced Biosciences, F38000, Grenoble, France
(2) Grenoble Alpes University, HP2 Laboratory, INSERM U1300, Grenoble Alpes University Hospital, Grenoble, France
(3) Institute of Transfusion Medicine and Transplant Engineering, Hannover Medical School, Hannover, Germany
(4) Pediatric Hematology and Oncology, Hannover Medical School, Hannover, Germany
(5) Univ Rouen Normandie, Inserm U1245, Normandie Univ, CHU Rouen, Department of Genetics, F-76000 Rouen, France
(6) Université Paris-Saclay, CEA, Centre National de Recherche en Génomique Humaine (CNRGH), 91057, Evry, France
(7) Halvorsen Center for Computational Oncology, Department of Epidemiology and Biostatistics, Memorial Sloan Kettering Cancer Center, New York, NY, USA
(8) Simons Center for Systems Biology, Institute for Advanced Study, Princeton, NJ, USA
† Deseased
*Equal contribution

Corresponding author: Pierre Hainaut, Institute for Advanced Biosciences, Univ. Grenoble Alpes, Inserm 1209, CNRS 5309, Site Santé, Allée des Alpes, F38700, La Tronche, France.

**Figure S1**

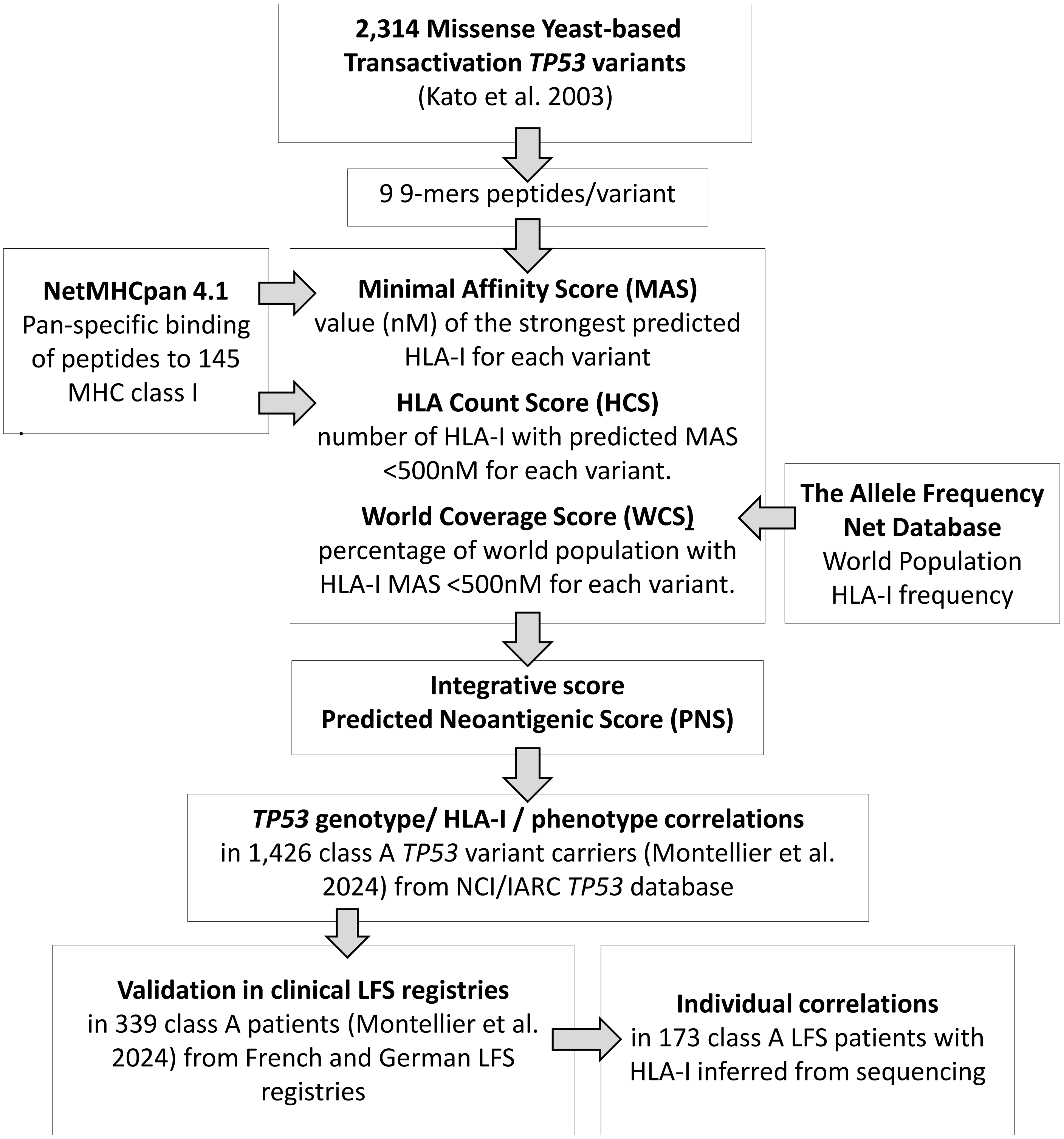
2,314 Missense Yeast-based Transactivation TP53 variants
(Kato et al. 2003)
9 9-mers peptides/variant
NetMHCpan 4.1
Pan-specific binding of peptides to 145 MHC class I
Minimal Affinity Score (MAS)
value (nM) of the strongest predicted HLA-I for each variant
HLA Count Score (HCS)
number of HLA-I with predicted MAS <500nM for each variant.
World Coverage Score (WCS)
percentage of world population with HLA-I MAS <500nM for each variant.
The Allele Frequency Net Database
World Population HLA-I frequency
Integrative score
Predicted Neoantigenic Score (PNS)
TP53 genotype/ HLA-I / phenotype correlations
in 1,426 class A TP53 variant carriers (Montellier et al. 2024) from NCI/IARC TP53 database
Validation in clinical LFS registries
in 339 class A patients (Montellier et al. 2024) from French and German LFS registries
Individual correlations
in 173 class A LFS patients with HLA-I inferred from sequencing

### Figure S2

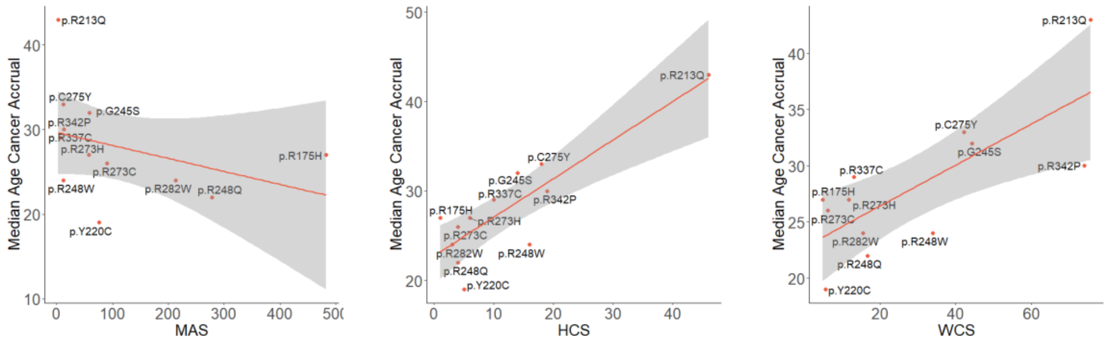


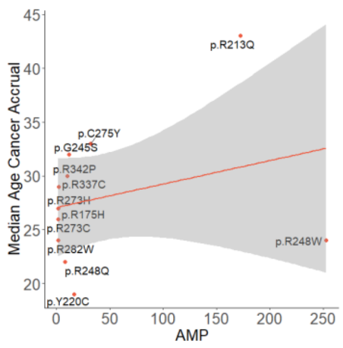


MAS spearman p=0.0276, rho = -0.63
HCS spearman p=0.01683, rho = 0.67
WCS spearman p=0.03367, rho = 0.61
AMS spearman p=0.4349, rho = 0.24

**Figure S3**

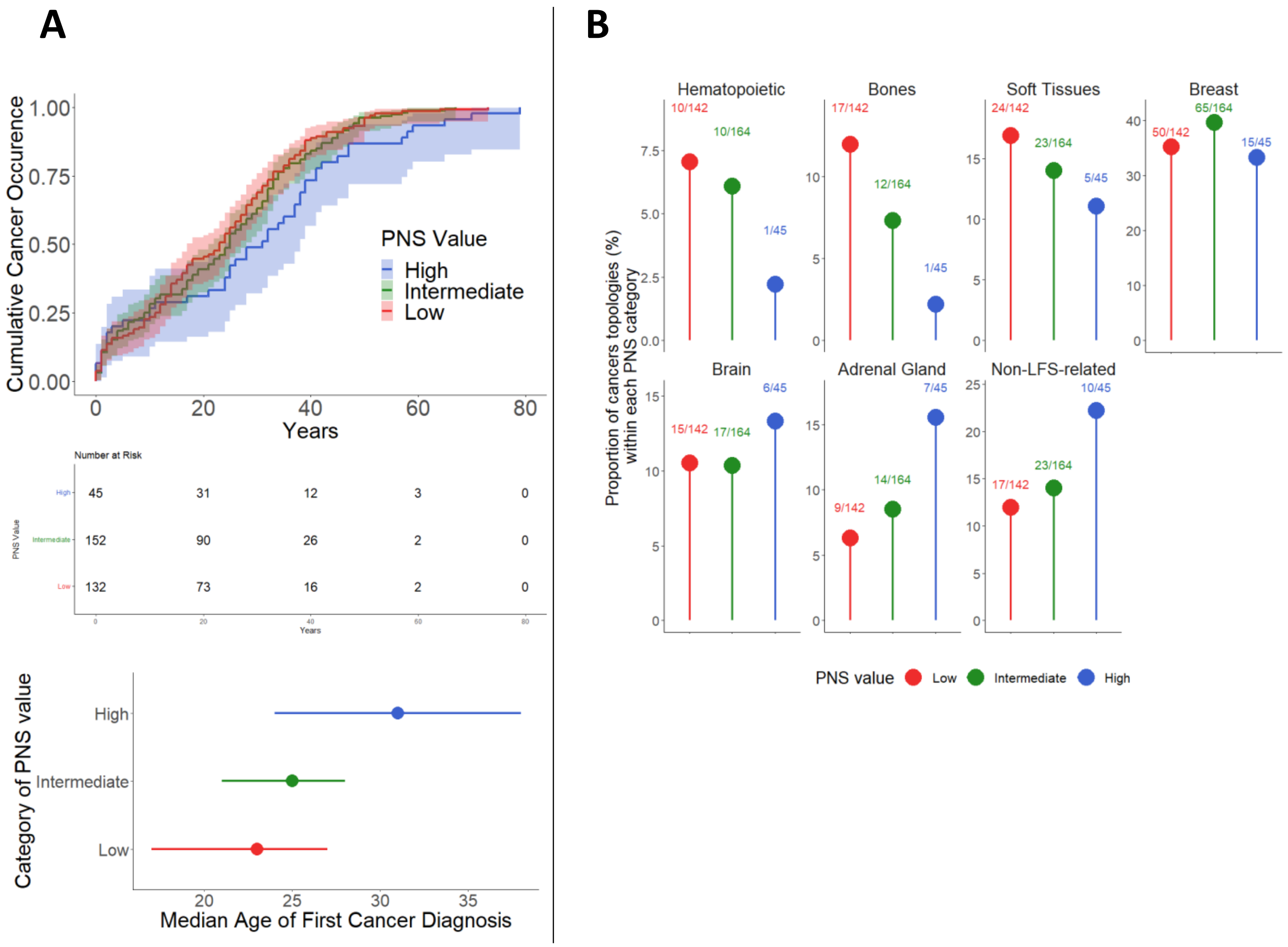
A
Cumulative Cancer Occurence
1.00
0.75
0.50
0.25
0.00
0
20
40
60
80
Years
PNS Value
High
Intermediate
Low
Number at Risk
PNS Value
High 45 31 12 3 0
Intermediate 152 90 26 2 0
Low 132 73 16 2 0
Years
Category of PNS value
High
Intermediate
Low
20 25 30 35
Median Age of First Cancer Diagnosis
B
Proportion of cancers topologies (%) within each PNS category
Hematopoietic
10/142
10/164
1/45
Bones
17/142
12/164
1/45
Soft Tissues
24/142
23/164
5/45
Breast
50/142
65/164
15/45
Brain
15/142
17/164
6/45
Adrenal Gland
9/142
14/164
7/45
Non-LFS-related
17/142
23/164
10/45
PNS value
Low
Intermediate
High

### Figure S4

A

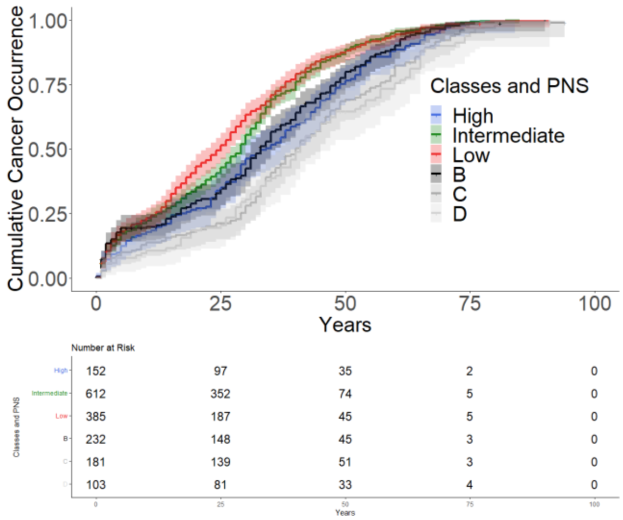


| Comparison | Adjusted_P_Value | Significance |
|---|---|---|
| High vs. Intermediate | 7.63e-03 | ** |
| Intermediate vs. Low | 1.00e+00 | NS |
| Low vs. High | 3.54e-03 | ** |
| High vs. B | 1.00e+00 | NS |
| High vs. C | 1.13e-01 | NS |
| High vs. D | 5.82e-02 | NS |
| Intermediate vs. B | 8.06e-03 | ** |
| Intermediate vs. C | 4.07e-11 | *** |
| Intermediate vs. D | 1.98e-08 | *** |
| Low vs. B | 4.56e-03 | ** |
| Low vs. C | 9.85e-11 | *** |
| Low vs. D | 4.76e-08 | *** |

B

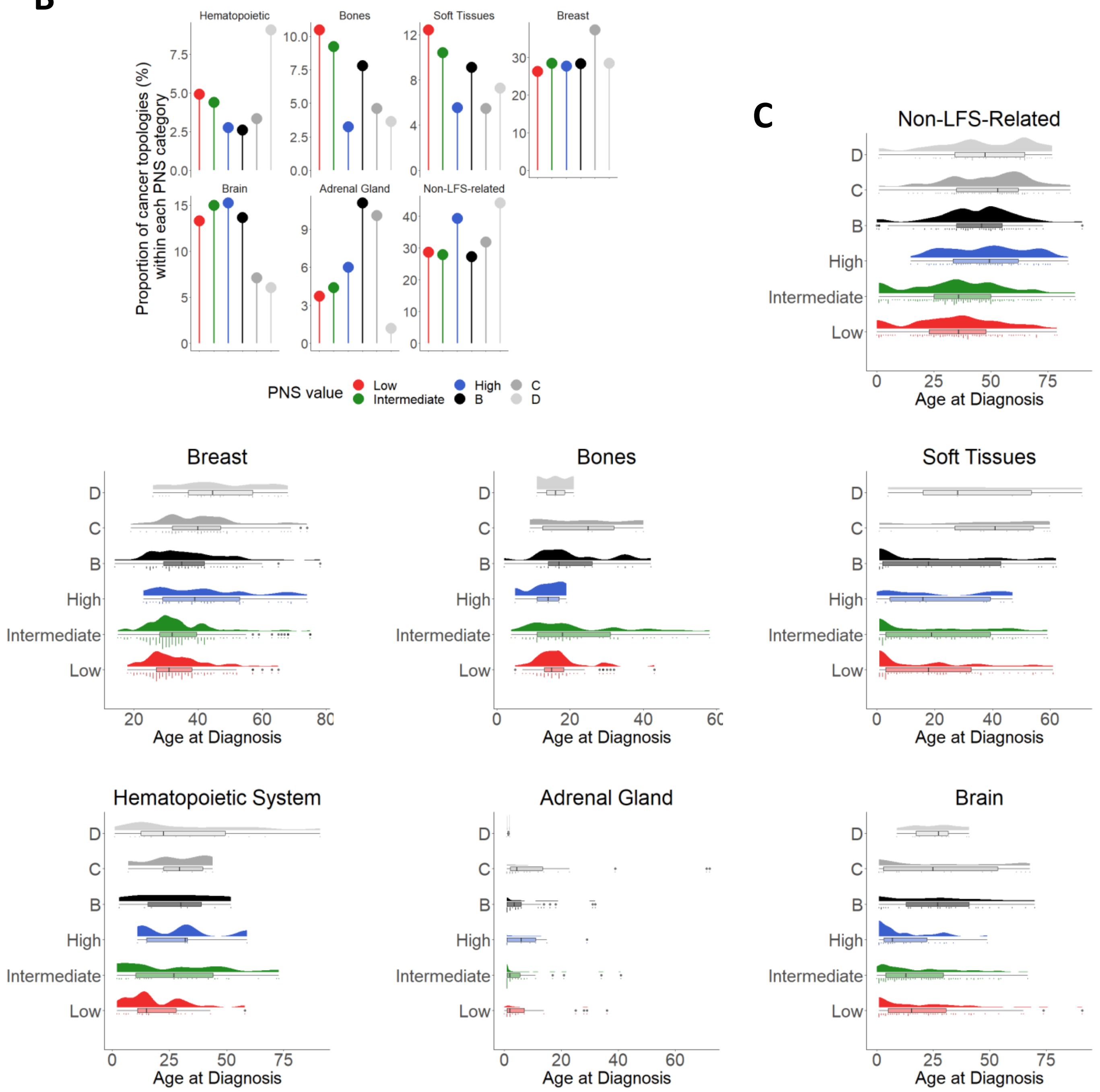
